# Association of Otolithic Integrity With Subjective and Functional Outcomes in Vestibular Rehabilitation: A Pilot Study

**DOI:** 10.64898/2026.04.01.26349994

**Authors:** Yunuen Hernández Cortés, Daniel Ramos Maldonado, Victoria Sosa Romo, Gómez-Coello Annel, Ivonne Calderón Leyva

**Author notes:** **Corresponding Author**: Daniel Ramos Maldonado, MD, Number: 5255411779988.

## Abstract

Variable recovery in vestibular rehabilitation underscores the need for objective biomarkers to identify patients at risk of poor clinical outcomes. This study aimed to establish proof of concept for a multidimensional prognostic framework using structural cervical vestibular evoked myogenic potential (cVEMP) and functional modified Clinical Test of Sensory Interaction on Balance (mCTSIB) markers to predict therapeutic success. This prospective cohort study was conducted at a tertiary rehabilitation center between June 2023 and May 2025. Participants were adults with peripheral vestibular disorders, including unilateral vestibular dysfunction, Meniere disease, or superior semicircular canal dehiscence. All participants underwent a customized five-session vestibular rehabilitation protocol.

Primary outcomes were subjective clinical success, defined as an 18-point reduction in Dizziness Handicap Inventory (DHI) score, and functional success, defined as a 3-point increase in Dynamic Gait Index score. Among 30 participants (mean age 60.8 years; 77% female), the rehabilitation protocol was associated with significant improvements in mean DHI (53.7 to 37.8; P = .003) and Dynamic Gait Index (19.5 to 22.1; P = .003) scores. While 83% of participants showed raw DHI improvement, only 37% achieved the 18-point minimal clinically important difference. Notably, no participants in the bilateral cVEMP absence group achieved subjective success, compared with 52.6% in the bilateral present group (P trend = .08). Multivariable logistic regression identified baseline DHI severity as an independent predictor of success (odds ratio, 1.05; 95% CI, 1.00–1.10; P = .04). Functional gait success was significantly correlated with baseline vestibular and visual preference ratios.

These findings suggest that baseline otolithic structural integrity is a primary determinant of subjective recovery. Bilateral structural loss may represent a “structural floor” where meaningful relief is physiologically limited despite functional gains. These results support a precision-based model using structural and sensory biomarkers to tailor rehabilitation.

**Trial Registration:** Mexican Institute of Rehabilitation (INR) Research Registry: INRLGII 62/22 PA URL: https://www.inr.gob.mx/Descargas/Transparencia/DatosA/2024-Anexo_estadistico_Investigacion.xlsx

## Introduction

Vestibular rehabilitation (VR) is the standard of care for patients with peripheral vestibular dysfunction, yet clinical outcomes remain highly variable [1, 2]. While most patients derive some benefit from exercise-based protocols, a significant subset fails to reach clinically meaningful thresholds for recovery. Current management strategies often rely on a “one-size-fits-all” approach because of a lack of robust prognostic markers that can accurately predict a patient’s capacity for compensation. Developing a precision medicine framework in VR requires an integrated understanding of how structural peripheral integrity and central functional plasticity interact to determine recovery.

The structural integrity of the otolithic system, specifically the saccule and its associated neural pathways, can be objectively quantified using cervical vestibular evoked myogenic potentials (cVEMPs) [3–5]. While cVEMPs are established tools for diagnosing specific pathologies [6, 7], their utility as a prognostic “floor”—a physiological limit below which subjective recovery may be precluded—remains poorly defined [8]. Furthermore, postural stability depends on the central nervous system’s ability to “reweight” multi-sensory inputs (visual, somatosensory, and vestibular), a process quantified by the modified Clinical Test of Sensory Interaction on Balance (mCTSIB) [9]. In clinical practice, a ratio of >0.8 is established as the threshold for normal integration, whereas a ratio of <0.8 signifies a specific deficit that necessitates targeted intervention [9].

Despite the availability of these structural and functional metrics, they are rarely synthesized into a unified prognostic model. Specifically, it remains unclear how absolute structural deficits (otolithic absence) versus functional integration deficits (sensory reweighting) independently contribute to subjective handicap, as measured by the Dizziness Handicap Inventory (DHI) [10, 11], and objective gait stability, measured by the Dynamic Gait Index (DGI) [12]. Moreover, clinical studies often overlook the “ceiling effect” of baseline scores, where patients with lower initial handicap have less measurable room for improvement, potentially masking the true efficacy of the intervention.

This proof-of-concept study aimed to establish a multidimensional prognostic framework for VR by evaluating otolithic integrity as a structural floor for subjective recovery. Our results demonstrate that this integrated framework successfully identifies rehabilitation-resistant phenotypes, transitioning clinical care from generalized protocols toward a precision-based model.

## Methods

Written informed consent was obtained from all participants before inclusion in the study. All procedures were conducted in accordance with the principles outlined in the Declaration of Helsinki. This study was approved by the Research Ethics Committee at the National Institute of Rehabilitation *Luis Guillermo Ibarra Ibarra* (approval number: INRLGII 62-22 PA).

### Study design and participant selection

Following protocol registration and approval from the Research Ethics Committee, a prospective longitudinal study was conducted from June 1^st^ 2023 to May 1^st^ 2025 in a tertiary referral center in Mexico. Participants were recruited based on a clinical diagnosis of peripheral or functional vestibular pathology, including unilateral vestibular dysfunction (n=15), Meniere disease (n=6), superior semicircular canal dehiscence syndrome (n=4), persistent postural-perceptual dizziness (n=4), and benign paroxysmal positional vertigo (n=1). Inclusion criteria were limited to: adults (aged ≥ 18 years) with a recorded baseline cVEMP, documented prerehabilitation, and postrehabilitation measurements for the DHI, DGI, and mCTSIB. Participants were required to complete 5 to 7 customized VR sessions. Exclusion criteria included incomplete data.

### Group classification and structural integrity

Participants were stratified into 3 groups based on baseline cVEMP status to represent a gradient of otolithic structural integrity: Group A (bilateral presence; n=19), Group B (unilateral absence or asymmetry ratio >40%; n=6), and Group C (bilateral absence; n=5).

### Primary outcomes and success thresholds

Therapeutic efficacy was defined by validated clinical success thresholds. Subjective success was defined as an 18-point reduction in the Dizziness Handicap Inventory (DHI) score, representing the Minimal Clinically Important Difference (MCID) [13]. Functional success was defined as a 3-point increase in the Dynamic Gait Index (DGI) score, representing the Minimal Detectable Change (MDC) [14]. Sensory integration was assessed via the modified Clinical Test of Sensory Interaction on Balance (mCTSIB); a baseline ratio of <0.8 was utilized as the cutoff for normal integration across the somatosensory (condition 2/condition 1), visual (condition 3/condition 1), vestibular (condition 4/condition 1), and visual preference (condition 3/condition 4) ratios [9].

### Statistical Analysis

Nonparametric methods were used given the cohort size (N=30). Continuous variables are presented as means (SDs) and categorical data as percentages. The global effectiveness of the rehabilitation protocol was assessed using the Wilcoxon signed-rank test. Bivariate associations between baseline cVEMP status and clinical success were evaluated using Fisher’s exact tests. Spearman’s rank correlation (*ρ*) was used to assess the relationship between baseline mCTSIB ratios and the magnitude of change in clinical scores.

A bi-dimensional prognostic model was constructed using multivariable logistic regression. To isolate the independent prognostic weight of structural and functional markers, models were adjusted for baseline symptom severity (prerehabilitation DHI and DGI scores). For instances of quasi-complete separation, specifically the 0% success rate in Group C, the analysis prioritized absolute success proportions and Fisher’s exact test results to preserve the clinical relationship between bilateral otolithic absence and rehabilitation resistance. A Cochran-Armitage trend analysis, approximated via logistic regression with ordinal coding for VEMP categories (0, bilateral present; 1, unilateral absence; 2, bilateral absence), was used to evaluate the dose-response relationship between deficit severity and recovery. Statistical significance was set at 2-tailed P < .05. Analysis was conducted using Python, version 3.10

## Results

### Cohort Characteristics and Baseline Integrity

The final study cohort included 30 participants (23 [77%] female; 7 [23%] male). The mean (SD) age of the total cohort was 60.8 (14.2) years, ranging from 57.0 (12.8) years in Group A to 69.0 (4.2) years in Group C. Baseline structural otolithic integrity, measured by cervical vestibular evoked myogenic potential (cVEMP) amplitude, varied across the three groups. In Group A, the mean (SD) right cVEMP amplitude was 83.618 ± 77.785 *μ*V with an asymmetry ratio of 20.816% ± 11.836%. Group B demonstrated a mean (SD) right cVEMP amplitude of 43.800 ± 21.131 *μ*V and a mean asymmetry ratio of 91.863% ± 19.931%. Group C participants had no cVEMP data at baseline (Table 1).

**Table 1:**
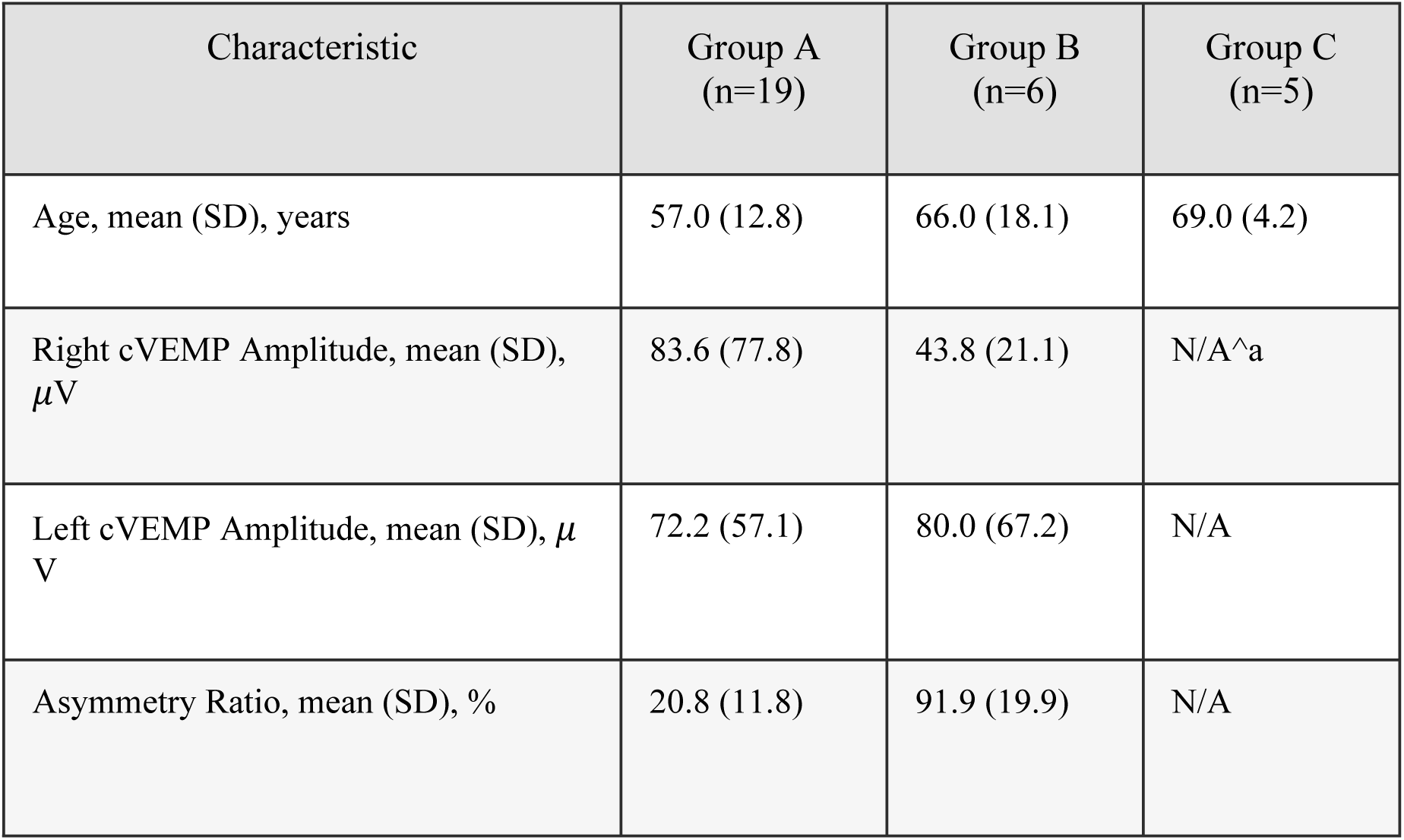
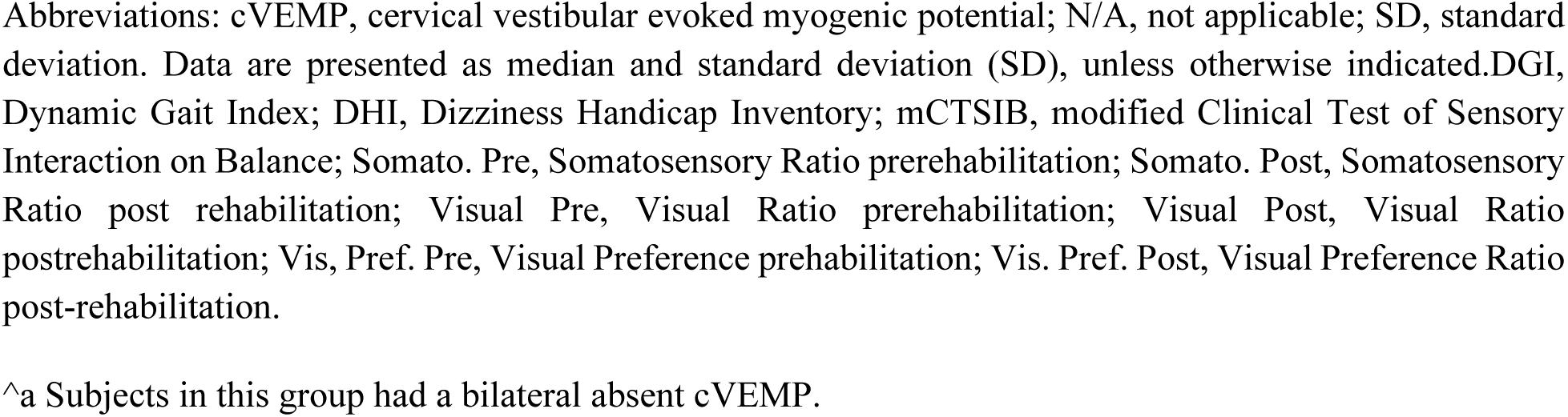
Baseline Demographic and Clinical Characteristics of 30 Participants.

| Characteristic | Group A<br>(n=19) | Group B<br>(n=6) | Group C<br>(n=5) |
| --- | --- | --- | --- |
| Age, mean (SD), years | 57.0 (12.8) | 66.0 (18.1) | 69.0 (4.2) |
| Right cVEMP Amplitude, mean (SD), $\mu\text{V}$ | 83.6 (77.8) | 43.8 (21.1) | N/A<sup>a</sup> |
| Left cVEMP Amplitude, mean (SD), $\mu\text{V}$ | 72.2 (57.1) | 80.0 (67.2) | N/A |
| Asymmetry Ratio, mean (SD), % | 20.8 (11.8) | 91.9 (19.9) | N/A |

Abbreviations: cVEMP, cervical vestibular evoked myogenic potential; N/A, not applicable; SD, standard deviation. Data are presented as median and standard deviation (SD), unless otherwise indicated. DGI, Dynamic Gait Index; DHI, Dizziness Handicap Inventory; mCTSIB, modified Clinical Test of Sensory Interaction on Balance; Somato. Pre, Somatosensory Ratio prerehabilitation; Somato. Post, Somatosensory Ratio post rehabilitation; Visual Pre, Visual Ratio prerehabilitation; Visual Post, Visual Ratio postrehabilitation; Vis. Pref. Pre, Visual Preference prerehabilitation; Vis. Pref. Post, Visual Preference Ratio post-rehabilitation.
<sup>a</sup> Subjects in this group had a bilateral absent cVEMP.

### Subjective and Functional Outcomes

Baseline and post-rehabilitation outcomes for the cohort are summarized in Table 2. At baseline, the mean (SD) Dizziness Handicap Inventory (DHI) scores were 53.7 (21.3) in Group A, 40.3 (29.0) in Group B, and 60.0 (27.9) in Group C. Following rehabilitation, mean DHI scores improved to 37.8 (21.7) in Group A and 29.7 (29.4) in Group B, while Group C scores increased to 62.0 (24.4). Functional performance, as measured by the Dynamic Gait Index (DGI), showed mean (SD) postrehabilitation improvements in Group A (22.1 [2.3]) and Group B (22.0 [2.1]) compared with Group C (16.8 [8.4]). Variations in modified mCTSIB ratios were observed across all groups between the prerehabilitation and postrehabilitation periods.

**Table 2.** Baseline and Post-Rehabilitation Subjective and Functional Characteristics of 30 Participants.

| Outcome Measure<sup>a</sup> | Group A (n=19) | Group B (n=6) | Group C (n=5) |
| --- | --- | --- | --- |
| Subjective (DHI) |  |  |  |
| Prerehabilitation | 53.7 (21.3) | 40.3 (29.0) | 60.0 (27.9) |
| Postrehabilitation | 37.8 (21.7) | 29.7 (29.4) | 62.0 (24.4) |

| Functional (DGI) |  |  |  |
| --- | --- | --- | --- |
| Prerehabilitation | 19.5 (3.0) | 18.7 (2.9) | 18.8 (5.5) |
| Postrehabilitation | 22.1 (2.3) | 22.0 (2.1) | 16.8 (8.4) |
| mCTSIB Ratios |  |  |  |
| Somatosensory (Pre) | 0.81 (0.32) | 0.93 (0.16) | 0.76 (0.39) |
| Somatosensory (Post) | 0.98 (0.08) | 1.00 (0.00) | 0.97 (0.08) |
| Visual (Pre) | 0.84 (0.33) | 0.97 (0.20) | 0.73 (0.31) |
| Visual (Post) | 0.97 (0.08) | 0.94 (0.14) | 0.77 (0.31) |
| Vestibular (Pre) | 0.76 (0.38) | 0.57 (0.40) | 0.39 (0.45) |
| Vestibular (Post) | 0.84 (0.26) | 0.90 (0.16) | 0.62 (0.52) |
| Visual Preference (Pre) | 1.39 (1.10) | 3.64 (3.62) | 3.68 (4.33) |
| Visual Preference (Post) | 1.39 (1.10) | 3.64 (3.62) | 3.68 (4.33) |

Abbreviations: SD, standard deviation; DHI, Dizziness Handicap Inventory; DGI, Dynamic Gait Index; mCTSIB, modified Clinical Test of Sensory Interaction on Balance; Somatosensory Pre, Somatosensory Ratio prerehabilitation; Somatosensory Post, Somatosensory Ratio post rehabilitation; Visual Pre, Visual Ratio prerehabilitation; Visual Post, Visual Ratio postrehabilitation; Vis, Pref. Pre, Visual Preference prerehabilitation; Vis. Pref. Post, Visual Preference Ratio post-rehabilitation.
<sup>a</sup>Data are presented as mean (SD).

### Treatment Outcomes by Baseline Otolithic Function

Analysis of treatment outcomes using the Wilcoxon signed-rank test identified distinct recovery patterns based on initial otolithic function (Table 3). Participants with preserved cVEMP function (Group A) showed improvements in DHI scores (mean [SD], 53.7 [21.3] to 37.8 [21.7]; P=.003), Dynamic Gait Index (DGI) scores (mean [SD], 19.5 [3.0] to 22.1 [2.3]; P=.003), and visual condition (P=.04). In the unilateral cVEMP absence cohort (Group B), significant improvements were observed in both DHI (mean [SD], 40.3 [29.1] to 29.7 [29.4]; P=.04) and DGI (mean [SD], 18.7 [2.9] to 22.0 [2.1]; P=.03) measures. Participants with bilateral cVEMP absence (Group C) did not demonstrate significant changes in any measured category. Mean (SD) DGI scores in Group C changed from 18.8 (5.5) to 16.8 (8.4) (P=.44), and DHI scores remained stable (60.0 [27.9] to 62.0 [24.4]; P=1.00).

**Table 3.**
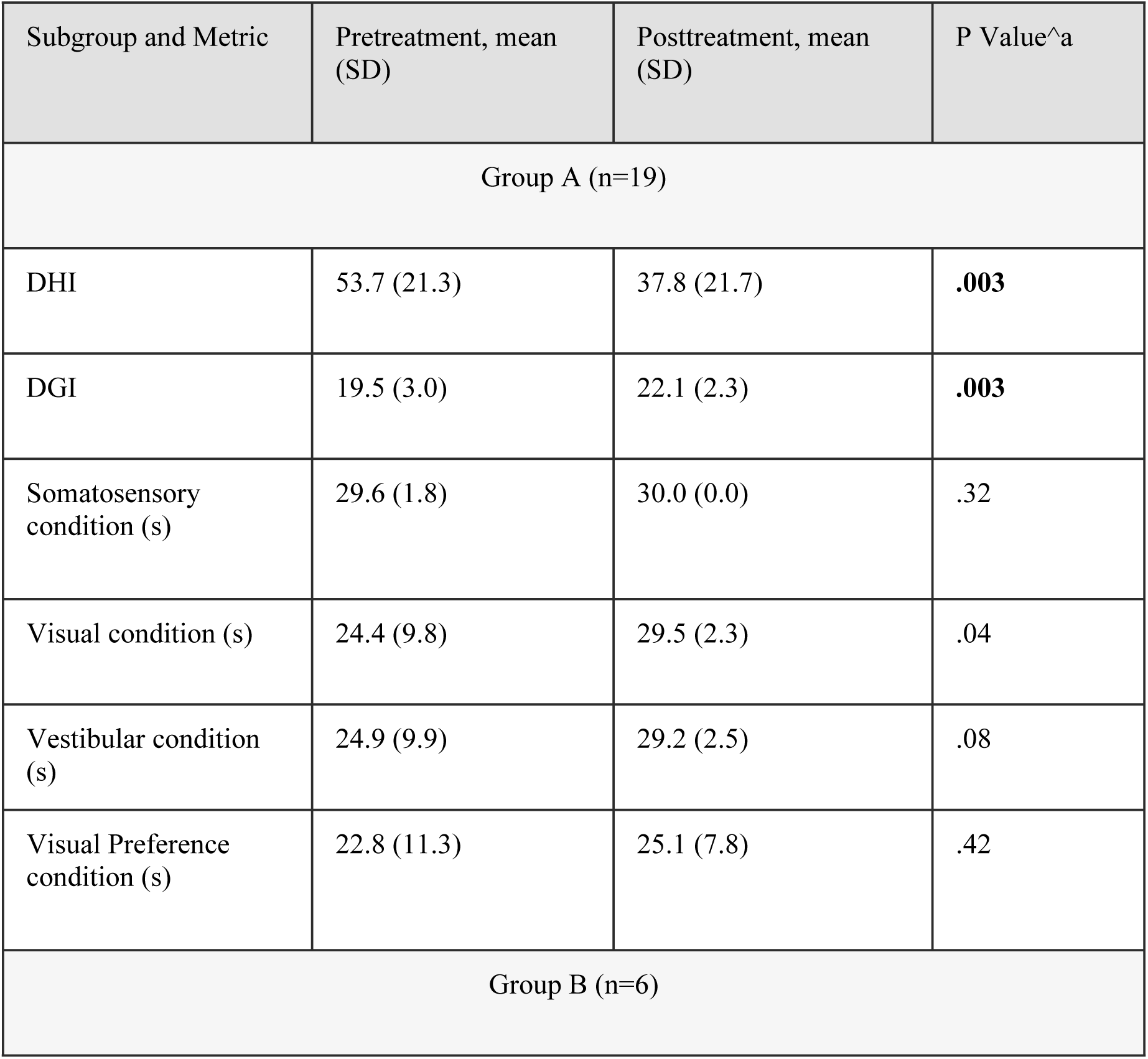
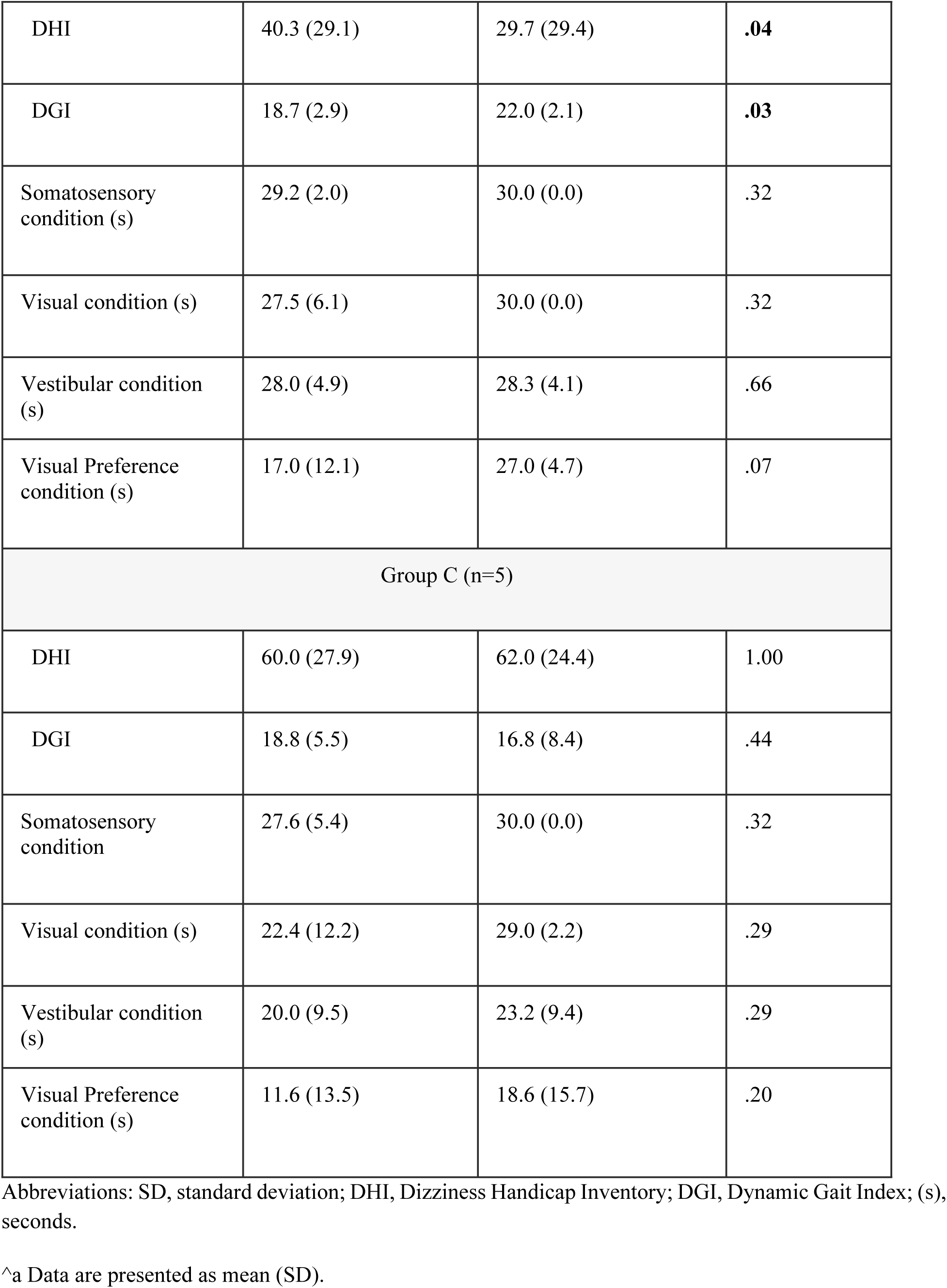
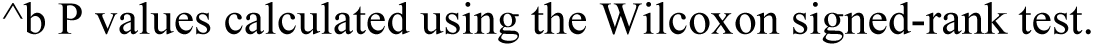
Pretreatment and Posttreatment Outcomes by cVEMP Subgroup among 30 Participants.

| Subgroup and Metric | Pretreatment, mean (SD) | Posttreatment, mean (SD) | P Value<sup>a</sup> |
| --- | --- | --- | --- |
| Group A (n=19) |  |  |  |
| DHI | 53.7 (21.3) | 37.8 (21.7) | **.003** |
| DGI | 19.5 (3.0) | 22.1 (2.3) | **.003** |
| Somatosensory condition (s) | 29.6 (1.8) | 30.0 (0.0) | .32 |
| Visual condition (s) | 24.4 (9.8) | 29.5 (2.3) | .04 |
| Vestibular condition (s) | 24.9 (9.9) | 29.2 (2.5) | .08 |
| Visual Preference condition (s) | 22.8 (11.3) | 25.1 (7.8) | .42 |
| Group B (n=6) |  |  |  |
| DHI | 40.3 (29.1) | 29.7 (29.4) | **.04** |
| DGI | 18.7 (2.9) | 22.0 (2.1) | **.03** |
| Somatosensory condition (s) | 29.2 (2.0) | 30.0 (0.0) | .32 |
| Visual condition (s) | 27.5 (6.1) | 30.0 (0.0) | .32 |
| Vestibular condition (s) | 28.0 (4.9) | 28.3 (4.1) | .66 |
| Visual Preference condition (s) | 17.0 (12.1) | 27.0 (4.7) | .07 |
| Group C (n=5) |  |  |  |
| DHI | 60.0 (27.9) | 62.0 (24.4) | 1.00 |
| DGI | 18.8 (5.5) | 16.8 (8.4) | .44 |
| Somatosensory condition | 27.6 (5.4) | 30.0 (0.0) | .32 |
| Visual condition (s) | 22.4 (12.2) | 29.0 (2.2) | .29 |
| Vestibular condition (s) | 20.0 (9.5) | 23.2 (9.4) | .29 |
| Visual Preference condition (s) | 11.6 (13.5) | 18.6 (15.7) | .20 |

Abbreviations: SD, standard deviation; DHI, Dizziness Handicap Inventory; DGI, Dynamic Gait Index; (s), seconds.
^a Data are presented as mean (SD).
<sup>a</sup> P values calculated using the Wilcoxon signed-rank test.

### Association of cVEMP Status With Clinical Success

Clinical success rates for subjective and functional outcomes, categorized by baseline cVEMP status, are summarized in Table 4. For the DHI, the clinical success rate (MCID ≥ 18) was 52.6% (10/19) in the bilateral present, 33.3% (2/6) in the unilateral absent group, and 0.0% (0/5) in the bilateral absent group. The odds ratio (OR) for failure to reach DHI success in the combined impaired group compared with the intact group was 5.00 (95% CI, 0.85-29.57; P=.12). Functional gait success (MDC ≥ 3) was achieved by 42.1% (8/19) of the intact group, 66.7% (4/6) of the unilateral absent group, and 20.0% (1/5) of the bilateral absent group. The OR for the combined impaired group reaching DGI success compared with the intact group was 0.87 (95% CI, 0.20-3.90; P> .99).

**Table 4:**
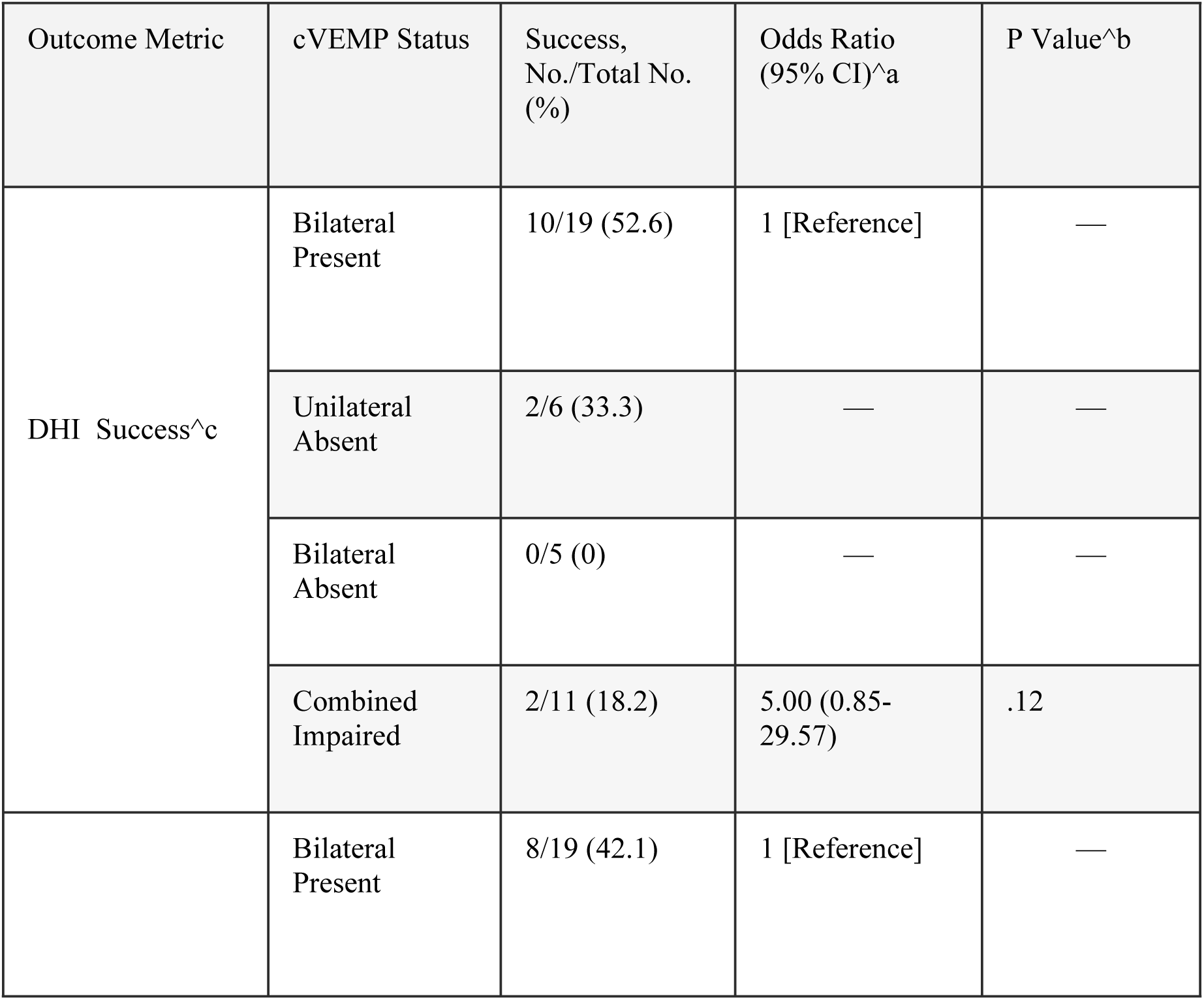
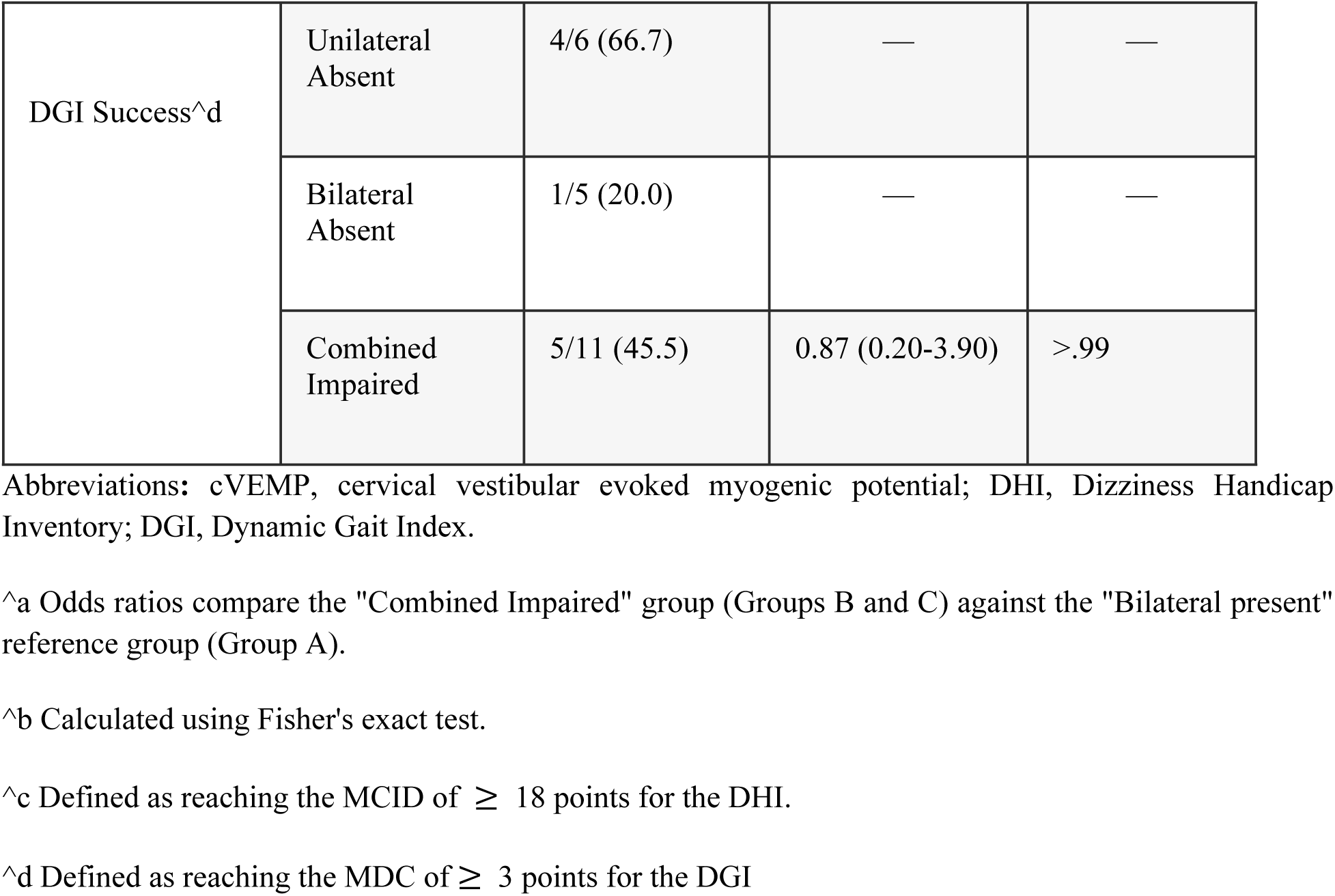
Association of Baseline cVEMP Status With Subjective and Functional Clinical Success.

### Correlation Between Baseline Sensory Integration and Recovery Outcomes

Spearman rank correlation coefficients for the associations between baseline sensory ratios and clinical recovery thresholds (DHI MCID, DGI MDC) are presented in Table 5. In Group B, a significant correlation was observed between the DGI MDC and both baseline vestibular ratios (*ρ*= -0.84; P = .04) and visual preference ratios (*ρ*= 0.84; P = .04). Correlations between baseline sensory ratios and clinical outcomes did not reach statistical significance in Groups A or C, or for the total cohort.

**Table 5.**
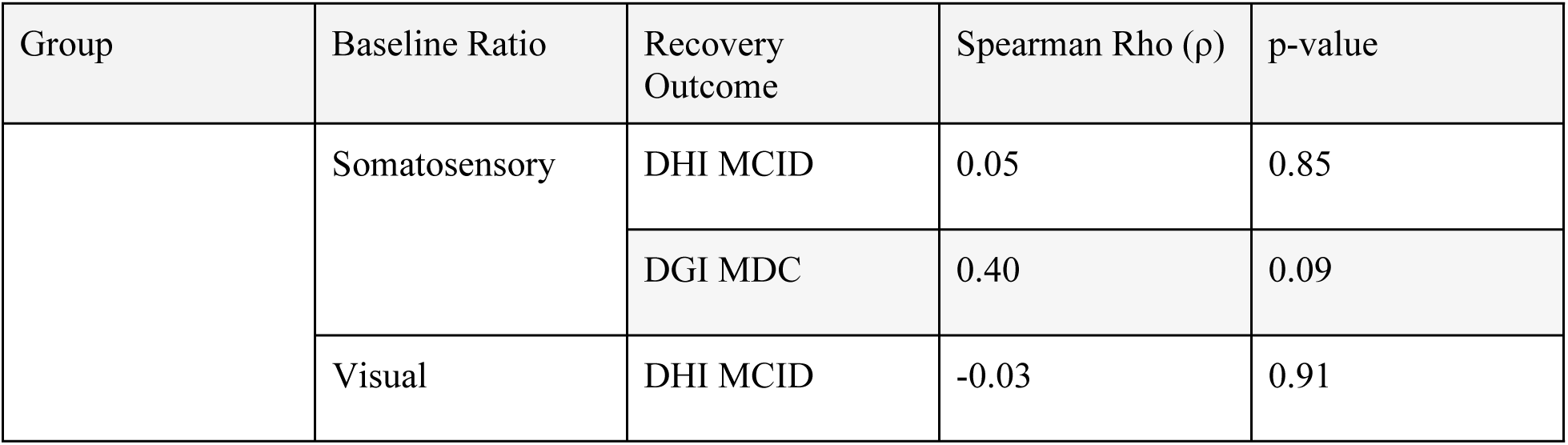
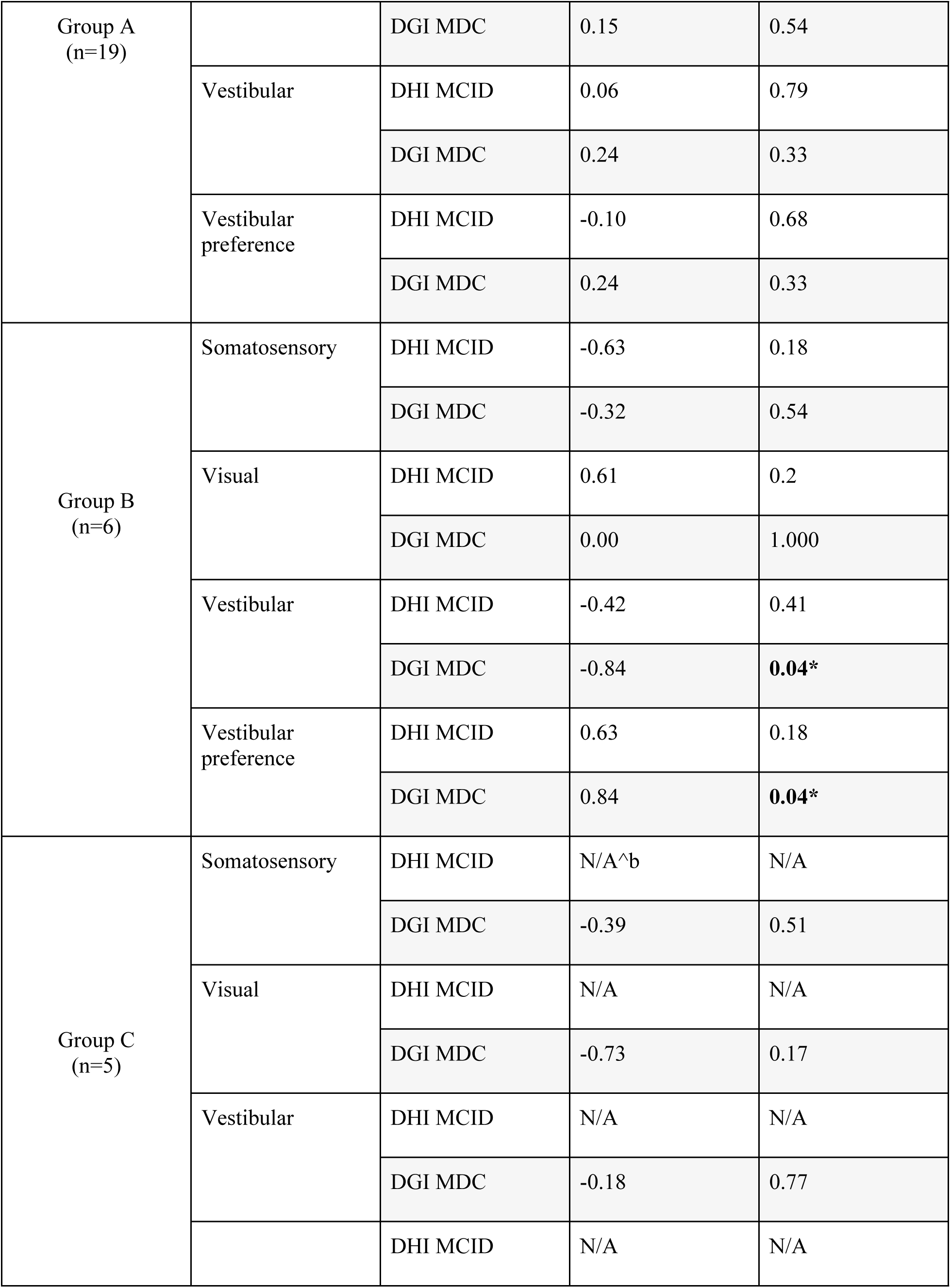
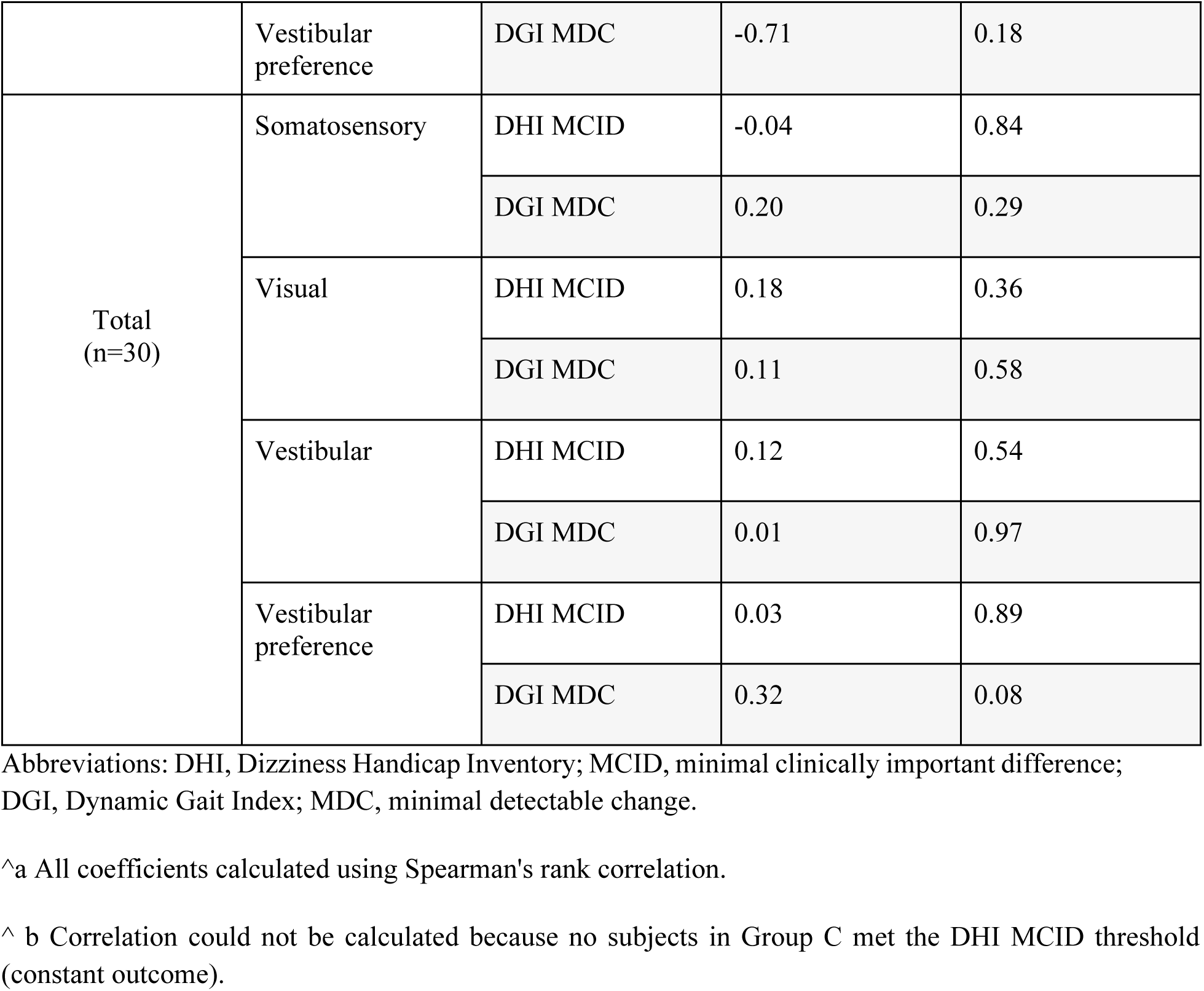
Spearman Rank Correlation Between Baseline Sensory Integration Ratios and Clinical Recovery Outcomes.

### Independent Predictors of Treatment Success

Multivariable logistic regression was used to identify independent predictors of treatment success (Table 6). Baseline Dizziness Handicap Inventory scores were significantly associated with DHI success (OR, 1.05; 95% CI, 1.00-1.10; P = .04); for every 1 point increase in the baseline DHI, the odds of success increase by 5.2%. cVEMP status and other baseline variables were not significant predictors of DHI success (P > .05). Within the cohort, 0 of 5 participants with bilateral cVEMP absence achieved DHI success compared with 47.4% (9/19) of participants with bilateral presence.

**Table 6:** Multivariable Logistic Regression Analysis of Factors Predicting Treatment Success in 30 Participants.

| Outcome and Predictor | Odds Ratio (95% CI) | P Value |
| --- | --- | --- |
| DHI Success<sup>a</sup> |  |  |
| Group B (Unilateral Absent) | 0.85 (0.09-8.49) | .89 |
| Group C (Bilateral Absent) | < 0.001 (0.00 to ∞)<sup>b</sup> | >.99 |
| Baseline DHI Score | 1.05 (1.00-1.10) | **.04** |
| DGI Success<sup>c</sup> |  |  |
| Group B (Unilateral Absent) | 2.74 (0.38-19.80) | .32 |
| Group C (Bilateral Absent) | 0.34 (0.03-3.73) | .38 |
| Baseline DHI Score | 1.00 (0.97-1.03) | .99 |

Abbreviations: DGI, Dynamic Gait Index; DHI, Dizziness Handicap Inventory; OR, odds ratio.
<sup>a</sup> Defined as reaching the minimal clinically important difference of $\geq 18$ points for the DHI.
<sup>b</sup> The zero percent success rate in Group C resulted in an indeterminate OR.
<sup>c</sup> Defined as reaching the minimal detectable change of $\geq 3$ points for the DGI.

For functional gait success (DGI), no baseline variables reached statistical significance in the multivariable model (P > .05). The OR for DGI success in participants with unilateral cVEMP absence was 2.74 (95% CI, 0.38-19.80; P = .32) compared with the intact reference group.

### Ordinal Trend Analysis of Vestibular Deficit Severity

Ordinal trend analysis demonstrated that with each progressive stage of severity of vestibular structural impairment (bilateral present, unilateral absent, and bilateral absent), the odds of achieving a significant DHI improvement decrease by approximately 72% (odds ratio [OR] per category increase, 0.28; 95% CI, 0.07-1.18; P *trend* = .08). While this trend approached statistical significance, it did not cross the 0.05 threshold. No significant trend was observed for Dynamic Gait Index (DGI) success (OR, 0.80; 95% CI, 0.31-2.10; P *trend* = .65) (Table 7).

**Table 7.** Ordinal Trend Analysis of Vestibular Structural Deficit Severity and Clinical Success among 30 Participants.

| Outcome | Odds Ratio (95% CI)<sup>a</sup> | P Value for Trend<sup>b</sup> |
| --- | --- | --- |
| DHI Success | 0.28 (0.07-1.18) | .08 |
| DGI Success | 0.80 (0.31-2.10) | .65 |

Abbreviations: CI, Confidence interval; DHI, Dizziness Handicap Inventory; DGI, Dynamic Gait Index;
<sup>a</sup> Data represent the odds of success per category increase in structural deficit severity (bilateral present, unilateral absent, and bilateral absent).
<sup>b</sup> Calculated using ordinal logistic regression

## Discussion

This proof-of-concept study establishes a multidimensional prognostic framework for VR based on the interplay between structural otolithic integrity, functional sensory integration, and baseline symptom severity. Our findings suggest that while functional gait recovery is driven by central adaptive plasticity, subjective relief from handicap is constrained by a “structural floor” dictated by the presence of at least one functional otolithic organ [8]. The most striking finding was the 0% success rate for achieving an MCID in the DHI among participants with bilateral cVEMP absence (Group C). This suggests that the total loss of sacculocollic integrity may represent a physiological limit to subjective recovery [8]. Although these patients showed improvements in raw scores, the absence of a gravitational reference frame provided by the otoliths appears to preclude the perception of meaningful relief. The observed trend toward decreased odds of DHI success as structural impairment progressed from normal to bilateral absence (OR = 0.28; 95% CI, 0.07–1.18; P = .08) supports a dose-response relationship between otolithic integrity and subjective prognosis [8]. A critical dissociation was observed between subjective handicap and objective functional recovery. Despite the “floor” effect in DHI scores, participants with unilateral deficits (Group B) achieved the highest rate of functional gait success (66.7%). This suggests that functional stabilization is driven principally by central adaptation rather than peripheral restoration [1, 5].

The predictive models indicate that baseline sensory integration, specifically, visual preference and vestibular ratios, serves as a primary marker for this plasticity. In Group B, the strong correlation between baseline visual preference and DGI recovery (*ρ* = 0.84; P = .04) highlights a mechanism of sensory substitution, where the central nervous system successfully “upweights” non-vestibular cues to restore physical balance even when the internal perception of dizziness persists [9]. Multivariable analysis identified baseline DHI scores as a significant independent predictor of success (P = .04), with higher initial handicap scores associated with a greater likelihood of reaching the 18-point MCID. This “therapeutic window” effect is common in clinical trials; however, our framework adds a necessary layer of precision by demonstrating that this window is effectively closed in the presence of bilateral structural loss. Conversely, functional gait recovery was limited by a ceiling effect; patients starting with high DGI scores had a lower probability of achieving the +3 point MDC [12]. This underscores the importance of adjusting for baseline status when evaluating rehabilitation efficacy to avoid underestimating the progress of more functional patients. These findings align with observations by Greeshma et al., who noted that structural deficits often persist even as clinical scores improve [8]. While 83% of the current cohort demonstrated a reduction in raw DHI scores, only 36.7% met the strict 18-point MCID threshold [13].3 This distinction is critical for precision medicine, as it separates statistical improvement from the meaningful clinical relief required for patient satisfaction.

### Limitations and Future Directions

This study has several limitations. The small cohort size (N = 30) limits the generalizability of the findings and the statistical power of the trend analyses. Validation in larger, etiology-specific groups is required to confirm the “structural floor” across different pathologies. Additionally, this study focused on the sacculocollic pathway via cVEMPs; future iterations of this prognostic framework should integrate oVEMP data to provide a more comprehensive utriculo-saccular profile. Finally, the reliance on the DHI MCID may not fully capture the quality-of-life improvements in patients with low initial handicap who nonetheless experience significant functional gains.

## Conclusions

Otolithic structural integrity, as measured by cervical vestibular evoked myogenic potentials, is a primary determinant of subjective clinical success in vestibular rehabilitation. The identification of a “structural floor” in patients with bilateral otolithic absence suggests that at least one functional otolithic organ is a physiological prerequisite for perceiving meaningful reductions in handicap. Despite these structural constraints on subjective relief, functional gait recovery remains achievable through central adaptive plasticity and somatosensory reweighting. These findings support the adoption of a bidimensional prognostic framework, incorporating objective structural biomarkers and functional sensory ratios, to enable a precision medicine approach. Such a framework allows for the early identification of rehabilitation-resistant phenotypes and the targeted escalation of care, including optokinetic stimulation or cognitive-behavioral therapy, for patients with significant structural barriers to standard recovery.

## Statements and Declarations

## Acknowledgments

Not applicable.

## Data Availability Statement

All relevant data underlying the findings described in this study are available as a de-identified dataset in the Supporting Information files

## Author contributions

Dr. Hernandez Cortés, Dr. Ramos Maldonado, Dr. Gómez-Coello, and Dr. Sosa Romo made substantial contributions to the conception and design of the study. Dr. Sosa Romo, Dr. Ramos Maldonado, Dr. Gómez-Coello, Dr. Calderón Leyva, and Dr. Hernandez Cortés contributed to data acquisition, analysis, and interpretation. Dr. Sosa Romo, Dr. Ramos Maldonado, Dr. Hernandez Cortés, and Dr. Gómez-Coello drafted the manuscript. Dr. Ramos Maldonado and Dr. Gómez-Coello critically revised the manuscript for important intellectual content. All authors approved the final version for publication and agree to be accountable for all aspects of the work.

## References

1. Charpiot A, Fath L, Perruisseau-Carrier J, et al. Rehabilitación vestibular. EMC - Otorrinolaringol. 2022;51(4):1–7. 10.1016/S1632-3475(22)47040-5

2. Smółka W, Smółka K, Markowski J, et al. The efficacy of vestibular rehabilitation in patients with chronic unilateral vestibular dysfunction. Int J Occup Med Environ Health. 2020;33(3):273–282. 10.13075/ijomeh.1896.01330

3. Rosengren SM, Welgampola MS, Taylor RL, et al. Vestibular evoked myogenic potentials in practice: Methods, pitfalls, and clinical applications. Clin Neurophysiol Pract. 2019;4:47–68. 10.1016/j.cnp.2019.01.005

4. Murofushi T. Clinical application of vestibular evoked myogenic potential (VEMP). Auris Nasus Larynx. 2016;43(4):367–376. 10.1016/j.anl.2015.12.006

5. Lee SH, Kim HJ, Lee SJ. Vestibular evoked myogenic potentials in assessing vestibular function: A clinical review. Otol Neurotol. 2022;43(2):169–177. 10.1097/MAO.0000000000003418

6. Agrawal Y, Bremova T, Kremmyda O, et al. Clinical testing of otolith function: Perceptual thresholds and myogenic potentials. J Assoc Res Otolaryngol. 2013;14(6):905–915. 10.1007/s10162-013-0416-x

7. Yang TH, Young YH. Clinical usefulness of vestibular-evoked myogenic potential testing - A review. J Formos Med Assoc. 2024. 10.1016/j.jfma.2024.11.008

8. Greeshma J, Kumar K, Ebenezer A. Comparison of VEMP and DHI in patients with peripheral vestibular lesions between pre- and post-vestibular rehabilitation. Int Tinnitus J. 2019;23(2):69–73. 10.5935/0946-5448.20190012

9. Shumway-Cook A, Horak FB. Assessing the attentional demands of balance tasks. J Mot Behav. 1986;18(4):435–446. 10.1080/00222895.1986.10735388

10. Koppelaar-van Eijsden HM, Schermer TR, Bruintjes TD. Measurement properties of the dizziness handicap inventory: A systematic review. Otol Neurotol. 2022;43(3):e282–97. 10.1097/MAO.0000000000003448

11. Zamyslowska-Szmytke E, Politanski P, Jozefowicz-Korczynska M. Dizziness Handicap Inventory in clinical evaluation of dizzy patients. Int J Environ Res Public Health. 2021;18(5):2210. 10.3390/ijerph18052210

12. Wrisley DM, Walker ML, Echternach JL, et al. Reliability and validity of the Dynamic Gait Index in individuals with vestibular disorders. Arch Phys Med Rehabil. 2003;84(10):1515–1521. 10.1016/S0003-9993(03)00274-0

13. Ahmed R, Elbaz A, Shoukry M. Evaluating the Dizziness Handicap Inventory in vestibular rehabilitation outcomes: A longitudinal study. J Vestib Res. 2020;30(4):247–254. 10.3233/VES-200696

14. Whitney SL, Marchetti GF, Scherer MR. The impact of vestibular dysfunction on the DHI in vestibular rehabilitation patients. Arch Phys Med Rehabil. 2006;87(11):1545–1550. 10.1016/j.apmr.2006.08.027

